# Rivaroxaban for patients with coronary dissection after drug-coated balloon angioplasty : A Randomized Controlled Trial

**DOI:** 10.1101/2024.01.04.24300877

**Authors:** Kaiyuan Liu, Quan Guo, Shi Qingbo, Ming Nie, Haosen Yu, Zhiwen Zhang, Muwei Li

**Author notes:** **Address for correspondence**: Dr Muwei Li, Department of Cardiology, Central China Fuwai Hospital of Zhengzhou University, No. 1 Fuwai Road, Zhengzhou, Henan Province, China. **E-mail:**. **Disclosures**: All authors have reported that they have no relationships relevant to the contents of this paper to disclose.

## Abstract

**Background:** After drug-coated balloon (DCB) angioplasty in patients with de novo coronary lesions, coronary dissections can result in the continuous presence of thrombi within the vessel wall, potentially leading to late luminal loss. Rivaroxaban, a novel oral anticoagulant, may mitigate lumen loss attributable to coronary dissection.

**Objectives:** This study was conducted to evaluate the effectiveness and safety of low-dose rivaroxaban in improving outcomes for patients with dissections following DCB intervention.

**Methods:** This trial was a prospective, randomized, controlled study including patients with acute coronary syndrome (ACS) who exhibited non-flow-limiting dissections after DCB angioplasty for de novo coronary lesions. The rivaroxaban group received a standard dual antiplatelet therapy (DAPT) regimen along with 2.5 mg of rivaroxaban twice daily for one month, reverting to DAPT thereafter. The control group was treated with standard DAPT for 6 months. All patients underwent coronary angiography at the 6-month mark. The primary endpoint involved late lumen loss in the target vessel at 6 months, and the primary safety endpoint consisted of bleeding events, as classified according to BARC criteria (types 1-3 or 5). This study has been registered with ClinicalTrials.gov, number NCT05750758.

**Results:** Out of 140 randomized patients, 137 (69 in the rivaroxaban group, 68 in the control group) completed the study. Low-dose rivaroxaban was associated with a significant reduction in late lumen loss compared to controls (−0.12 ± 0.56 mm versus 0.14 ± 0.37 mm, P = 0.002). The rivaroxaban group experienced more minor bleeding events (BARC 1-2), though this was not statistically significant (P = 0.059). Clinical outcomes were similar between groups (P = 0.354).

**Conclusions:** This study demonstrates that the combination of rivaroxaban with dual antiplatelet therapy shows promise in reducing stenosis and enhancing late lumen expansion in lesions with dissections post-DCB intervention.

## Introduction

The drug-coated balloon (DCB) represents a type of medical device coated with anti-vascular intimal hyperplasia drugs. Upon inflation at the diseased vessel site, the balloon contacts the vessel wall’s endothelium. The drug is then rapidly released onto the endothelium, disrupting and stretching the vessel wall, inhibiting smooth muscle cell proliferation, and thus reducing restenosis rates without necessitating metallic stent implantation. Evidence has established the safety and efficacy of DCB treatment in various conditions, including in-stent restenosis^1^, and bifurcation lesions^2^. There is also an increasing body of evidence supporting the use of DCB in the treatment of de novo lesions, especially in patients with small vessel disease, high bleeding risk, and myocardial infarction^3–6^. Also, evidence is growing regarding the efficacy of a DCB-only strategy in treating de novo lesions in large (≥2.75 mm) coronary arteries^7–9^.

The utilization of DCB-only therapy is progressively being adopted in the management of de novo lesions. However, mechanically, DCB function similarly to conventional balloons, and consequently, they inherit some of the primary limitations associated with these devices following angioplasty, specifically coronary dissection and acute recoil.

Dissection is a double-edged sword. Dissections might enhance the contact area between the vessel wall and antiproliferative drugs, promoting drug absorption. Conversely, severe dissections may also lead to the persistence of thrombi within the vessel wall, contributing to late lumen loss post-thrombosis organization or even directly leading to complications like acute myocardial infarction^10^. These risks highlight the need for complementary therapeutic strategies to mitigate complications and enhance overall treatment efficacy.

In this context, rivaroxaban, a novel oral anticoagulant, emerges as a promising approach. Studies like ATLAS ACS 2-TIMI 51 and COMPASS have already underscored the efficacy of rivaroxaban in reducing major adverse cardiovascular events (MACE) in coronary heart disease^11–13^. This backdrop sets the stage for exploring rivaroxaban’s role in patients with coronary dissections post DCB angioplasty.

Taking into consideration the context outlined above, we have formulated this trial with the primary objective of assessing the effects of rivaroxaban for ACS patients with coronary dissection after drug-coated balloon angioplasty. To date, no studies have been conducted in this specific area. We hypothesize that rivaroxaban would improve clinical benefits in patients who retain dissections post-DCB treatment and aim to test this hypothesis in a prospective, randomized, controlled trial.

## Materials and Methods

### Trial design

The trial followed a prospective, randomized design. It complied with the Declaration of Helsinki and was approved by the New Business and Technology Ethics Committee of Fuwai Central China Cardiovascular Hospital, Zhengzhou University (Zhengzhou, China). We followed the Consolidated Standards of Reporting Trials (CONSORT) checklist^14^ to report this study (Supplemental file 1: CONSORT checklist). All participants signed an informed consent form. This study is registered with ClinicalTrials.gov, number NCT05750758.

### Participants

Eligible participants were those: (1) Age≥18 years; (2) Acute coronary syndrome (ACS) patients meeting the criteria for percutaneous coronary intervention (PCI); (3) DCB used for de novo target lesions during PCI; (4) Non-flow limited dissection. Exclusion criteria: (1) Age≥60 years; (2) Long-term oral anticoagulant treatment; (3) High bleeding risk (meeting the major or minor criteria for high bleeding risk according to Academic Research Consortium)^15^; (4) Non-de novo vascular lesions; (5)Severe calcified lesions; (6) Contraindication to antiplatelet and anticoagulant therapy; (7) Allergy to heparin or contrast agent; (8) Planned elective surgery; (9) Expected life span < 1 year; (10) Need for stent bail out due to intra-procedural complications.

### Procedural Technique

The procedure followed the consensus guidelines from international and Asian experts on drug-coated balloons^3, 16^. Preoperatively, patients received standard dual antiplatelet therapy (DAPT), and intraoperatively, heparin (70-100U/kg) was administered. Coronary angiography was performed following nitroglycerin-induced dilation to obtain optimal target lesion images. Predilation strategies were determined by the operator based on lesion characteristics. Intra-vascular ultrasound (IVUS) and rotational atherectomy were employed as needed. In cases of flow-limiting dissection after predilation, PCI using a drug-eluting stent (DES) was recommended without using a DCB. DCBs used were SeQuent (B. Braun, Germany) and adhered to standard operating protocols. The DCB was inflated for 30 to 60 s at nominal pressure. After using a DCB, a final assessment was undertaken at least 5 minutes after administering a bolus of intracoronary vasodilator, in order to catch acute vessel recoil. In the event of vessel recoil, bailout stent implantation was considered. TIMI flow and dissection status were recorded immediately post-procedure.

Patients with any residual coronary dissection after DCB use entered the current analysis. It is our habit not to stent coronary dissections of type A to C (National Heart, Lung, and Blood Institute [NHBLI] classification system for intimal tears)^17^ with Thrombolysis In Myocardial Infarction (TIMI) flow grade 3.

### Randomization and interventions

Randomization (1:1 ratio) was performed by an independent statistician using a computer-generated list. The rivaroxaban group received a standard DAPT(Aspirin 100mg QD + Ticagrelor 90mg BID) regimen along with 2.5 mg of rivaroxaban twice daily for one month, reverting to DAPT for 5 months thereafter. The control group was treated with standard DAPT for 6 months. Angiographic follow-up was conducted at 6 months post-procedure.

### Endpoints

The primary endpoint was LLL at 6 months, and the secondary endpoint was target vessel failure (TVF, composed of cardiac death, target vessel myocardial infarction, target vessel revascularization, and target vessel thrombosis)^18^. The main safety endpoint was bleeding as defined by the BARC 1-3 or 5 criteria^19^.

### Follow Up and Data Analysis

Telephone follow-up assessments were conducted at 1, 3, and 6 months postoperatively to document cardiovascular medication usage, clinical status, and adverse events, with a specific emphasis on bleeding events. All bleeding and ischemic events were independently adjudicated by a Clinical Events Committee^18, 19^. Subsequently, all patients underwent angiographic follow-up with quantitative coronary assessment after 6 months. The assessment of quantitative analysis for angiographic data was conducted by an independent core laboratory (Key Laboratory of Coronary Artery Imaging Medicine of Henan Province, Zhengzhou, China), utilizing the CAAS research system (Version 5.10, Pie Medical Imaging B.V., Maastricht, The Netherlands). The laboratory remained uninformed about the data grouping during the analysis phase, adhering to a blinded methodology to ensure impartiality in the evaluation process. Analyses were conducted on angiograms with identical projections before, after interventions, and at the angiographic follow-up. All measurements were performed on cineangiograms recorded after 200 mg of intracoronary nitroglycerin administration. Identical projections were used for each comparison. Measurements were taken for minimum luminal diameter (MLD), maximal diameter stenosis (DS%), reference vessel diameter (RVD), lesion length, binary restenosis, and persistence of dissection (NHLBI classification), and late luminal enlargement (LLE) or late luminal loss (LLL). LLL was defined as a decrease in the minimum luminal diameter at the follow-up compared to the immediate post-procedural measurement. The calculation method involved subtracting the immediate post-procedural MLD from the follow-up MLD. A positive result (i.e., >0) indicated LLE, while a negative result (i.e., <0) indicated LLL. Measurements encompassed the entire treated segment plus 5 mm proximally and distally. Binary restenosis was defined as stenosis of at least 50% of the luminal diameter at angiographic follow-up.

### Statistical analysis

The present trial’s sample size was determined from our pilot study, where a 0.20 mm difference in the 6-month LLL change from baseline was observed across two randomized groups. Group sample sizes of 66 per group achieve 90% power to detect an LLL mean difference of 0.2 mm, a standard deviation of 0.4 mm in each group, and a significance level (alpha) of 0.05, using a two-sided two-sample equal-variance t-test. Considering the anticipated dropout rate, each group was planned to include 70 patients.

Statistical analysis was performed using SPSS software (version 25.0; SPSS Inc., Armonk, NY). Categorical variables are presented as the number of cases and percentages. Comparisons between groups were performed using the chi-square test. Continuous variables conforming to normal distributions are presented as the mean± standard deviation. Comparisons between groups were performed using the unpaired Student’s t-test. Continuous variables not conforming to normal distributions are expressed as median (interquartile ranges). Between-group comparisons were processed using the Mann–Whitney U test.

## Results

From May 2022 to January 2023, we recruited 235 patients with ACS who were hospitalized in Fuwai Central China Cardiovascular Hospital. Among them, two patients were lost to follow-up after randomization, one participant refused to return to hospital for endpoint assessment (Figure 1). A total of 137 patients (69 in the rivaroxaban group and 68 in the control group) completed the treatment and endpoint assessments, of which 119 (90.15%) were male, with an average age of 50.72±6.49 years. All patients successfully underwent PCI using a DCB, with type A-C dissections (as classified by the NHLBI). All participants used the SeQuent DCB manufactured by B. Braun, Germany. No stents were used for any lesions during hospitalization. Baseline clinical characteristics, shown in Table 1, revealed that 32% had diabetes mellitus, and clinical indications were unstable angina (62%), and non-ST-segment elevation myocardial infarction (38%). The baseline characteristics were comparable between the two randomized groups.

**Figure 1.**
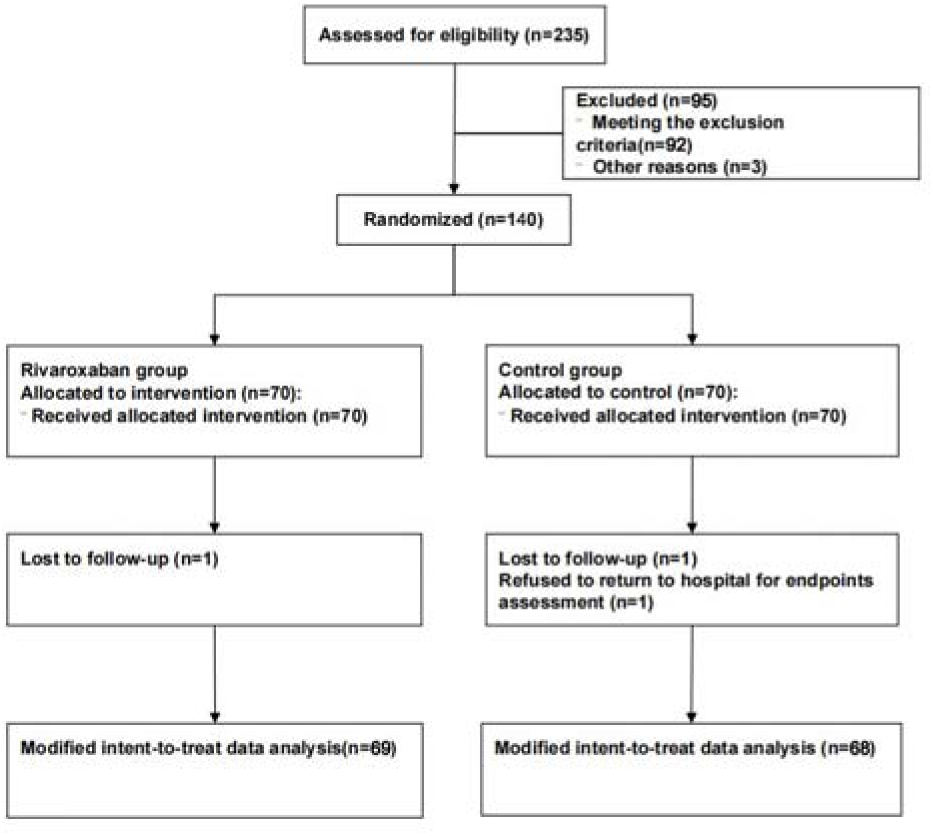
flow diagram for the study

**Table1.** Baseline characteristics of the participants.

|  | Rivaroxaban group(n=69) | Control group(n=68) | P |
| --- | --- | --- | --- |
| Male | 59(85.5%) | 60(88.2%) | 0.637 |
| Age , years | 50.12±6.45 | 51.34±6.52 | 0.272 |
| Hypertension | 30(43.5%) | 21 (30.9%) | 0.127 |
| Hypercholesterolemia | 36(52.2%) | 30 (44.1%) | 0.345 |
| Diabetes mellitus | 9(13%) | 16(23.5%) | 0.112 |
| History of smoking | 40(58%) | 36(52.9%) | 0.554 |
| History of drinking | 30(43.5%) | 29(42.6%) | 0.922 |
| BMI | 30.89±10.93 | 32.10±11.66 | 0.535 |
| Prior MI | 7(10.1%) | 4(5.8%) | 0.359 |
| Prior PCI | 9(13%) | 9(13.2%) | 0.974 |
| Haemoglobin(g/dL) | 138.52±13.46 | 140.53±15.29 | 0.416 |
| Creatinine clearance<br>(mL/min) | 94.38±14.52 | 89.908±12.678 | 0.069 |
| Clinical presentation |  |  |  |
| Unstable angina | 44(64%) | 41(60%) |  |
| Non-ST-segment elevation<br>MI | 25(36%) | 27(40%) | 0.675 |

Values are mean±SD, median (interquartile ranges, 25th–75th), or n (%). MI, myocardial infarction; PCI, percutaneous coronary intervention; CAD, coronary artery disease.

Table 2 presents baseline angiographic and procedural characteristics, demonstrating comparability in RVD, lesion length, MLD, operation time, the use of scoring balloons for predilation. We found that the majority of the dissections were of Type A(69.3%), and the distribution of dissection types was similar in both groups, with no significant differences being observed.

**Table 2:**
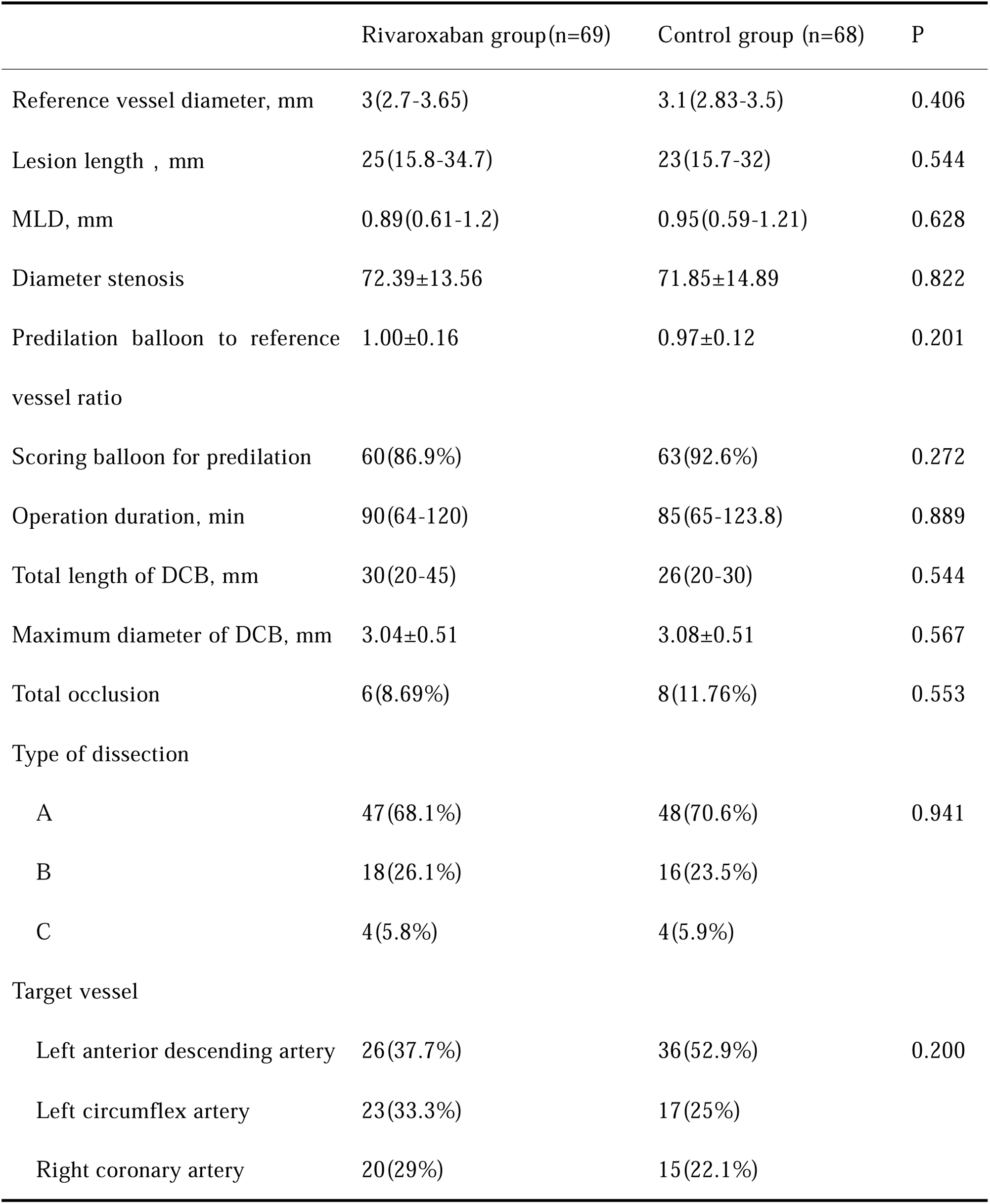
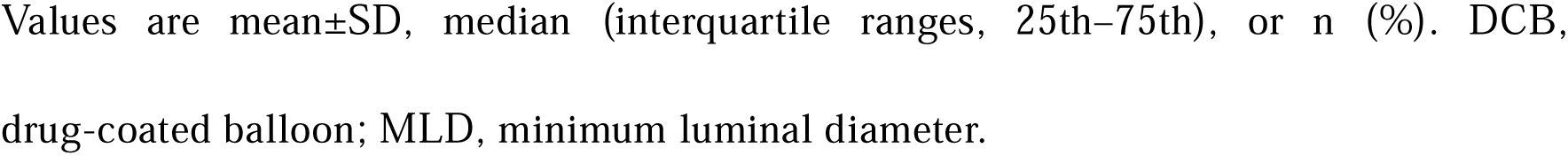
Angiographic and Procedural Characteristics.

|  | Rivaroxaban group(n=69) | Control group (n=68) | P |
| --- | --- | --- | --- |
| Reference vessel diameter, mm | 3(2.7-3.65) | 3.1(2.83-3.5) | 0.406 |
| Lesion length , mm | 25(15.8-34.7) | 23(15.7-32) | 0.544 |
| MLD, mm | 0.89(0.61-1.2) | 0.95(0.59-1.21) | 0.628 |
| Diameter stenosis | 72.39±13.56 | 71.85±14.89 | 0.822 |
| Predilation balloon to reference vessel ratio | 1.00±0.16 | 0.97±0.12 | 0.201 |
| Scoring balloon for predilation | 60(86.9%) | 63(92.6%) | 0.272 |
| Operation duration, min | 90(64-120) | 85(65-123.8) | 0.889 |
| Total length of DCB, mm | 30(20-45) | 26(20-30) | 0.544 |
| Maximum diameter of DCB, mm | 3.04±0.51 | 3.08±0.51 | 0.567 |
| Total occlusion | 6(8.69%) | 8(11.76%) | 0.553 |
| Type of dissection |  |  |  |
| A | 47(68.1%) | 48(70.6%) | 0.941 |
| B | 18(26.1%) | 16(23.5%) |  |
| C | 4(5.8%) | 4(5.9%) |  |
| Target vessel |  |  |  |
| Left anterior descending artery | 26(37.7%) | 36(52.9%) | 0.200 |
| Left circumflex artery | 23(33.3%) | 17(25%) |  |
| Right coronary artery | 20(29%) | 15(22.1%) |  |

Values are mean ± SD, median (interquartile ranges, 25th–75th), or n (%). DCB, drug-coated balloon; MLD, minimum luminal diameter.

Post-operative and follow-up stenosis analysis can be seen in Table 3. MLD at post-procedure was comparable in both groups, being 2.20 mm (IQR: 1.78-2.50) in the rivaroxaban group versus 2.23 mm (IQR: 1.7-2.53) in the control group (P = 0.690). The acute lumen gain was also comparable in both groups (P = 0.879). Patients underwent scheduled angiographic follow-up with quantitative coronary assessment after 190 days (IQR: 173-223 days). At follow-up, there was a noticeable difference in MLD between the rivaroxaban group and the control group (2.37 mm, IQR: 1.96-2.78 versus 2.16 mm, IQR: 1.75-2.41; P = 0.010). Meanwhile, a significant difference in net lumen gain between the rivaroxaban group and the control group was observed (1.43 ± 0.81 mm versus 1.16 ± 0.59 mm, P = 0.024). The primary endpoint of LLL showed a significant difference between the rivaroxaban group and the control group (−0.12 ± 0.56 mm versus 0.14 ± 0.37 mm, P = 0.002) (Figure 4). Of note, this negative LLL rather represents a late lumen enlargement in rivaroxaban group.

**Table 3:** Analysis of Lesion Stenosis at Post-Procedure and Follow-Up.

|  | Rivaroxaban group(n=69) | Control group (n=68) | P |
| --- | --- | --- | --- |
| MLD at post-procedure,<br>mm | 2.20(1.78-2.50) | 2.23(1.70-2.53) | 0.690 |
| Diameter stenosis at<br>post-procedure | 28.94±15.32 | 30.95±13.28 | 0.413 |
| Acute lumen gain, mm | 1.31±0.54 | 1.29±0.56 | 0.879 |
| MLD at follow-up, mm | 2.37(1.96-2.78) | 2.16(1.75-2.41) | 0.010 |
| Diameter stenosis at<br>follow-up | 25.46 ±23.50 | 35.12±16.54 | 0.006 |
| Late lumen Loss, mm | -0.12±0.56 | 0.14±0.37 | 0.002 |
| Binary restenosis, mm | 1(1.4%) | 2(2.9%) | 0.989 |
| Net lumen gain, mm | 1.43 ±0.81 | 1.16 ±0.59 | 0.024 |
| Dissection at follow-up |  |  |  |
| None | 65(94.2%) | 62(91.2%) | 0.725 |
| A | 4(5.8%) | 6(8.8%) |  |
| B | 0 | 0 |  |
| C | 0 | 0 |  |

Values are mean±SD, median (interquartile ranges, 25th–75th), or n (%). MLD, minimum luminal diameter.

During follow-up angiography, over 90% of the dissections in both groups healed completely, with 10 cases of persistent but uncomplicated dissections (Figure 2). There was no significant difference in the healing of dissections between the two groups.

**Figure 2.**
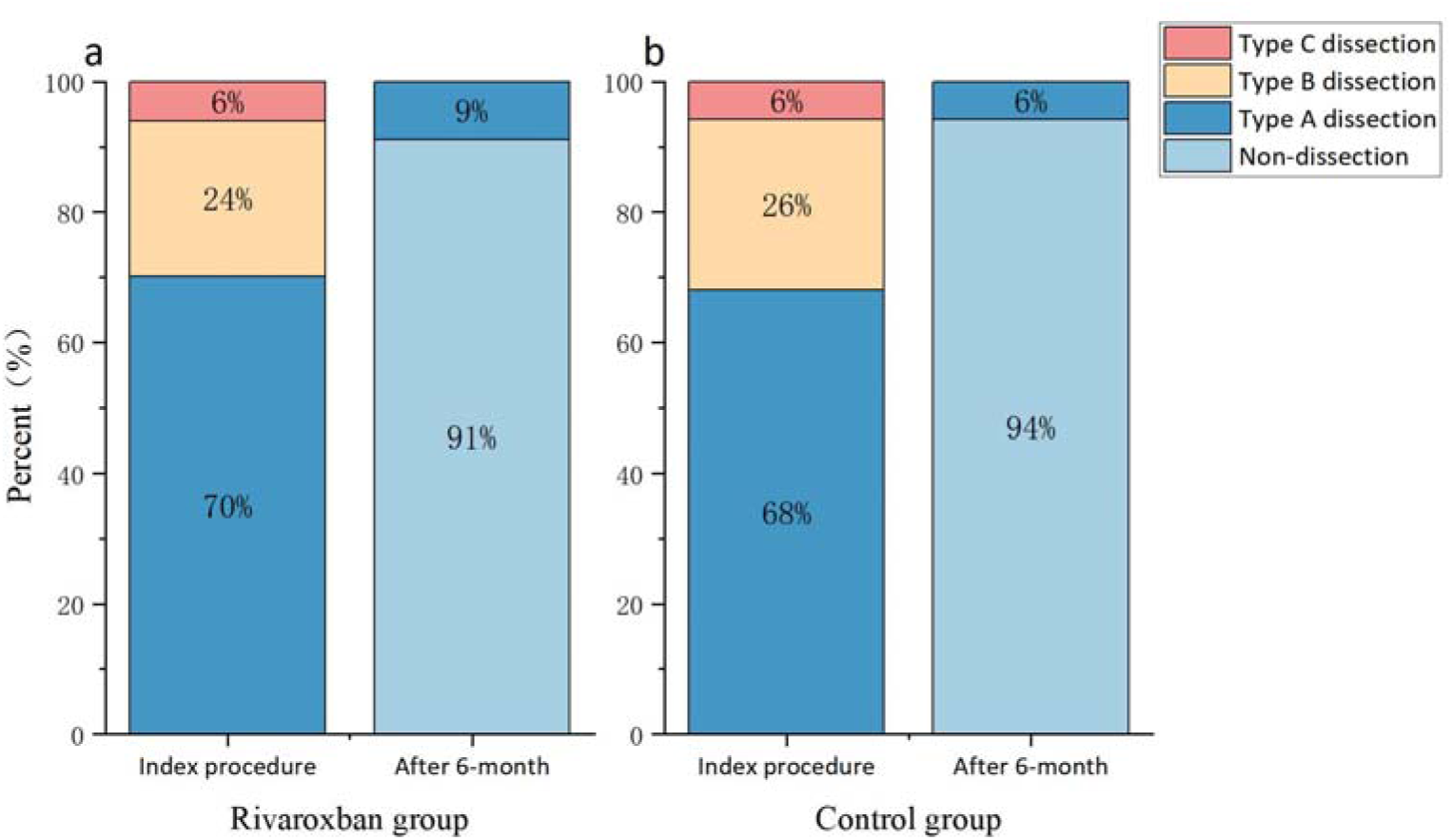
The Fate of Dissections After DCB Angioplasty Figure shows what happened to dissections at 6-month angiography: Over 90% of the dissections remaining post-DCB in both groups healed completely, with 10 cases of persistent but uncomplicated dissections.

Table 4 indicates more BARC type 1-2 bleeding events in the rivaroxaban group, suggesting minor bleeding events without an increased probability of major bleeding. One patient in the rivaroxaban group had a type 3a BARC bleeding event, resulting in a decrease in hemoglobin to 63 g/L and requiring blood transfusion. TVF occurred in 1 patient (1.5%) in the rivaroxaban group and in 4 patients (5.8%) in the control group, primarily due to TVR (Table 5, P = 0.354).

**Table 4:**
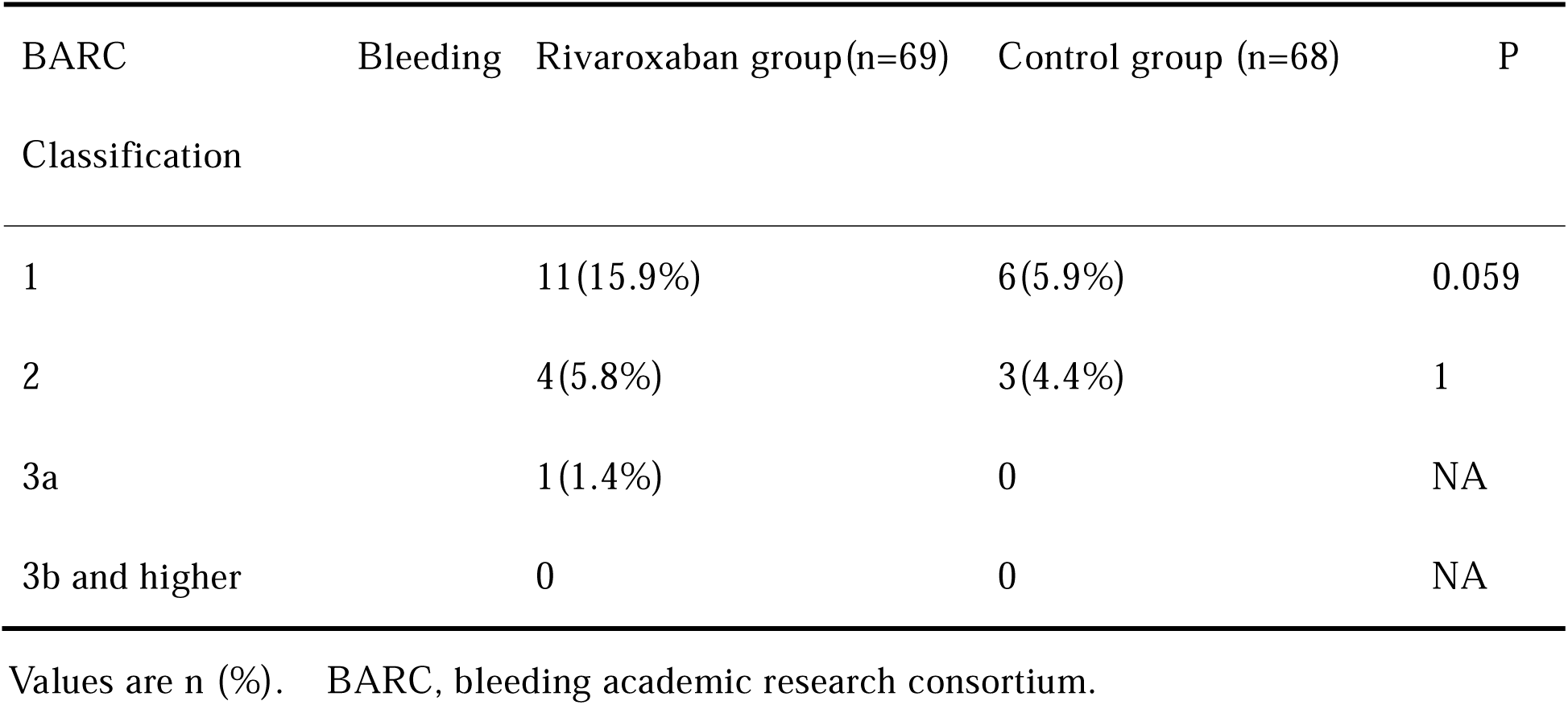
Bleeding Event Endpoints.

**Table 5:** Clinical Outcomes.

|  | Rivaroxaban group(n=69) | Control group (n=68) | P |
| --- | --- | --- | --- |
| Cardiac death | 0 | 0 | NA |
| Target vessel myocardial infarction | 0 | 0 | NA |
| Target vessel revascularization | 1(1.5%) | 4(5.8%) | 0.354 |
| Target vessel thrombosis | 0 | 0 | NA |
| Target vessel failure | 1(1.35%) | 4(5.8%) | 0.354 |

Values are n (%). Target vessel failure consisted of cardiac death, target vessel myocardial infarction, target vessel revascularization, and target vessel thrombosis.

## Discussion

This prospective study explored the impact of administering a low-dose rivaroxaban regimen for one months on imaging outcomes and clinical events in ACS patients with non-flow-limiting dissections following DCB intervention for de novo lesions. The key findings of this study are as follows: 1) The oral administration of low-dose rivaroxaban is associated with significant reduction in late lumen loss;2) Irrespective of rivaroxaban use, dissections tend to heal almost completely.

During the early era of balloon angioplasty (BA), the issue of vessel restenosis posed a significant challenge, often attributed to inadequate predilatation. Increasing balloon diameter was an attempt to improve suboptimal predilatation, but it concurrently raised the probability of dissection occurrence. Early experiences revealed that, despite a very low occurrence of thrombotic events and an acceptable rate of restenosis (12%), 36.7% of dissections following BA were still visible at 6-month angiographic follow-up^20^. In the case of DES edge dissections post-percutaneous coronary intervention, studies have demonstrated that dissections, particularly those with a smaller effective lumen area, are associated with target lesion revascularization during 1-year follow-up post-DES implantation^21^. However, in the DCB era, research by Cortese, et al. identified that dissections post-DCB intervention did not significantly correlate with restenosis or adverse events. Furthermore, a significant majority of dissections (94%) exhibited complete healing during follow-up angiography, irrespective of the dissection type^22^. These findings confirm the safety of non-flow-limiting dissections post-DCB intervention and underscore the potential of DCB in facilitating dissection healing.

In the DCB era, the presence of non-flow limiting dissections after DCB intervention was generally considered safe. In the third International DCB Consensus Report, the criteria for dissection management shifted from necessitating stent rescue for Type C-F dissections to those deemed as flow-limiting. Studies, including those by Yamamoto and Sogabe, have even suggested that dissections might predict late lumen enlargement^23^. Potential mechanisms involve dissections increasing the contact area between the drug-coated balloon and the vessel, thus facilitating better utilization of anti-proliferative agents like paclitaxel to inhibit intimal hyperplasia and promote vessel expansion. Furthermore, the disruption of the intima or media by dissections, influenced by hemodynamics, might contribute to further vessel dilation.

As a novel oral anticoagulant, rivaroxaban is the first direct factor Xa inhibitor, selectively targeting the active site of factor Xa. In addition to inhibiting the coagulation cascade, rivaroxaban also exerts antiplatelet effects by inhibiting platelet activation, which is mediated by FXa activation of the PAR-1 receptor, and by modulating PAR-1 and PAR-2 pathways involved in the inflammatory response^24, 25^. Multiple studies have shown that low-dose rivaroxaban therapy in coronary heart disease is linked with lower rates of MACE, albeit with an elevated bleeding risk^11–13^. Considering this, we established stringent exclusion criteria to omit patients at a relatively higher risk of bleeding from the study. Our observations indicated a marginally higher incidence of BARC 1-2 bleeding events (minor bleeding) in the rivaroxaban group compared to the control group, yet without an increase in major bleeding risk. However, large-scale studies with a larger sample size and longer follow-up time are needed to determine this result

In the setting of a spontaneous coronary section, although debatable, there are reports that antiplatelet can worsen outcomes. There are hypotheses that antithrombotic therapy can prolong the dissection by increasing the hematoma.^26^ But, the pathophysiological mechanisms and the affected patient population of spontaneous coronary artery dissection (SCAD) differ significantly from the context of our study. Most SCAD patients are females, and there may be increased vasa vasorum density, fibromuscular dysplasia, and other issues, leading to a higher risk of bleeding and different outcomes of administrating antithrombotic drugs.^27^

In Cortese, et al.’s study, about one-third of the lesions treated with DCB for de novo lesions exhibited dissections, yet the majority of these dissections (94%) had healed completely during follow-up coronary angiography. These results are comparable to those in the present study, in which dissections healed completely in 92.7% (Figure 2). In the control group, lesions with dissections post-DCB treatment showed late lumen loss during follow-up angiography, measured at 0.14±0.37 mm, comparable to the earlier study (0.14±0.28 mm)^22^. However, the rivaroxaban group generally exhibited late lumen enlargement (Figure 3), with a late luminal enlargement of 0.12±0.56 mm. Compared to the control group, a significant increase in lumen diameter was observed (P = 0.01) (Table 3). Net lumen gain was also notably larger in the rivaroxaban group (P = 0.024). These results indicate that rivaroxaban significantly improved both the degree of stenosis and the late luminal gain in these lesions. Previous research observed late lumen enlargement post-DCB intervention may be associated with the reattachment and healing of the dissection flap, and the regression of the dissection flap^28^. Reattachment and healing of the dissection flap were defined as the dissection flap protruding into the vessel lumen becoming reattached to the tunica media and healing. The regression of the dissection flap was defined as an obvious partial tissue loss in the dissection flap at follow-up, as compared to the post-PCI state. They discovered that the frequency of dissection flap regression was significantly higher in the LLE group (40.4% vs. 14.6%, p < 0.01). These observations suggest that promoting flap regression during the dissection occurrence and healing process might lead to greater late lumen expansion. This necessitates the use of robust antithrombotic therapy to maintain blood flow, thus playing a crucial role in the interaction between dissections and vessels, and in turn, facilitating lumen enlargement. Rivaroxaban, with its anticoagulant and anti-inflammatory properties, maintained robust antithrombotic activity during the acute phase of dissection formation, especially when combined with dual antiplatelet therapy. This approach correspondingly promoted maximal lumen expansion. As observed in this study, combining rivaroxaban with dual antiplatelet therapy improved stenosis severity and facilitated late lumen expansion in patients with dissections post-DCB intervention. This approach has the potential to enhance coronary blood flow, improve myocardial perfusion, reduce cardiac event risk. Meanwhile, in the control group, a higher incidence of TVF was observed, though it did not reach statistical significance, underscoring the need for further large-scale studies for confirmation.

**Figure 3.**
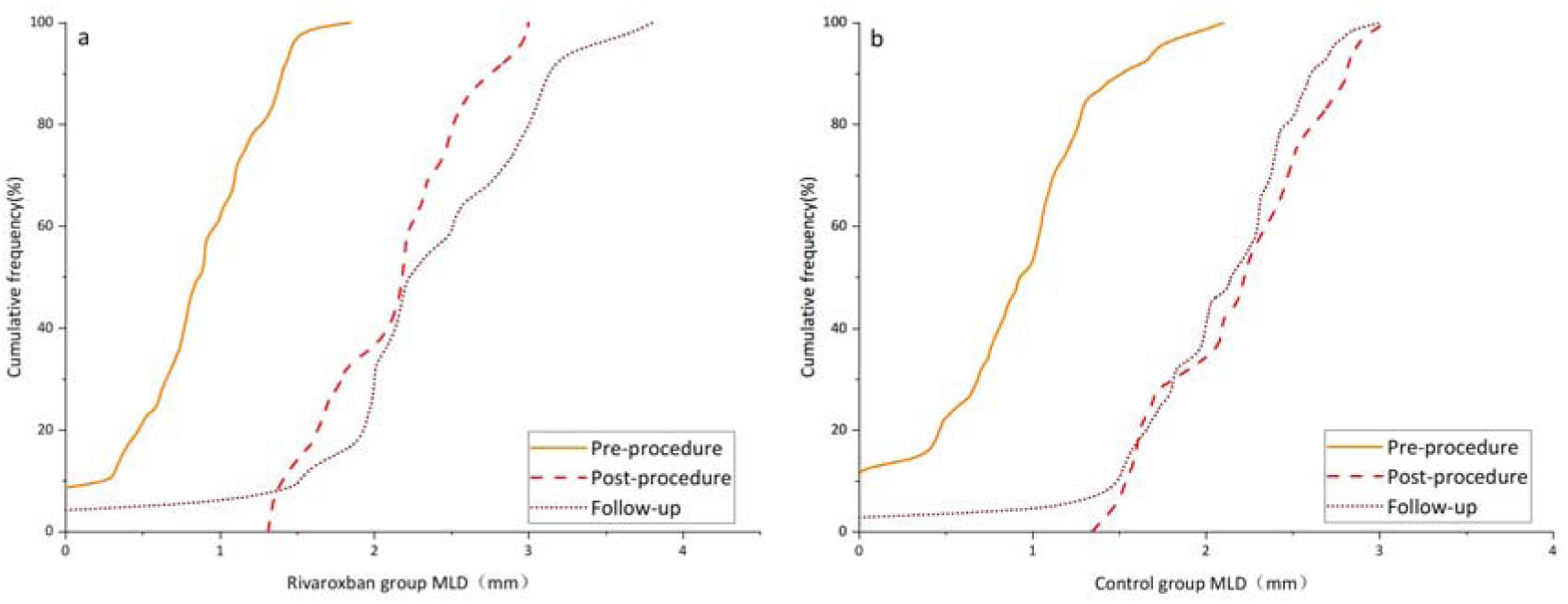
Cumulative frequency distribution curves of minimal lumen diameter of rivaroxaban group (a) and Control group (b) before procedure, after procedure, and at follow-up.

**Figure 4.**
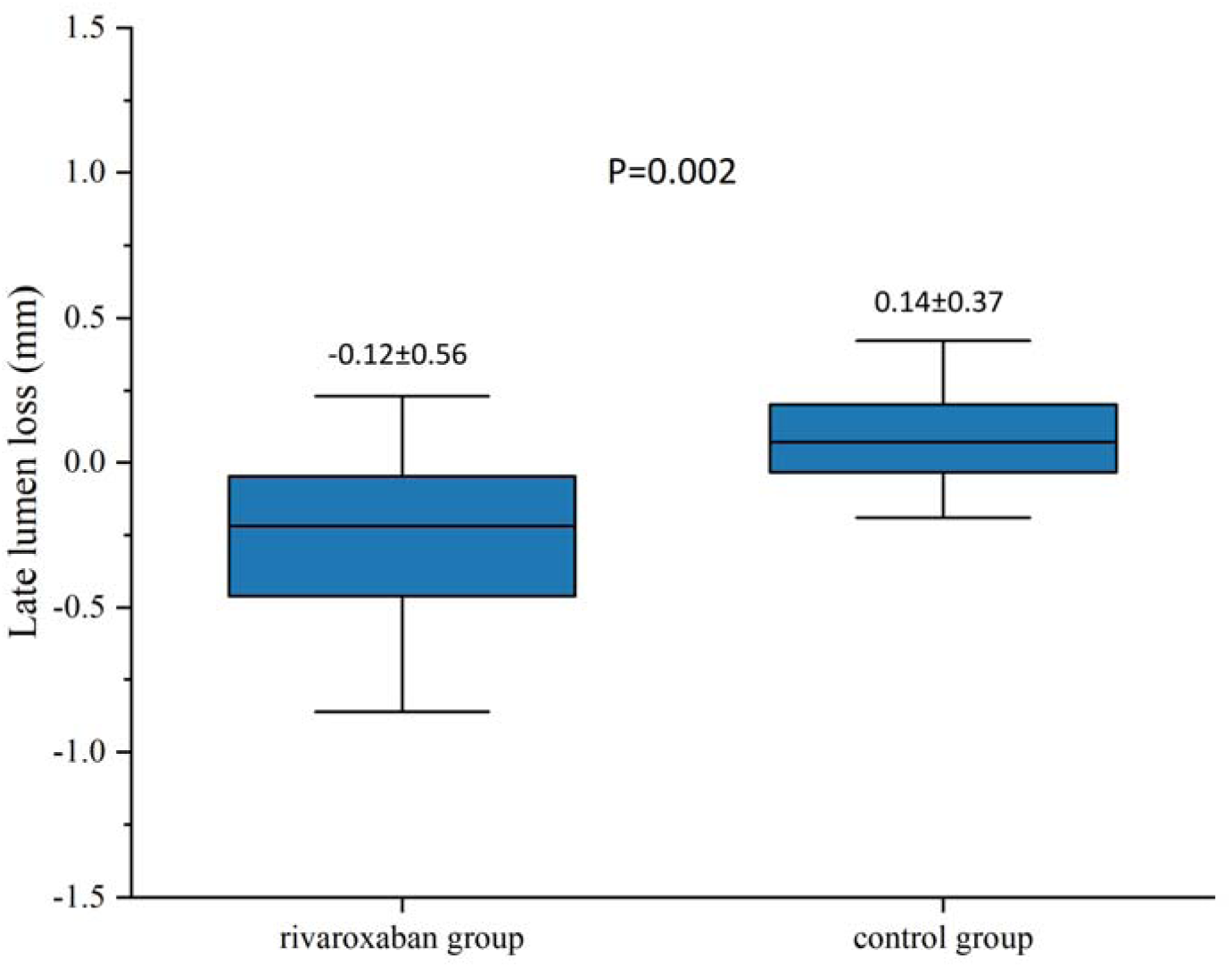
Comparison of late lumen loss

### Study limitations

Several limitations should be acknowledged in our study. Firstly, observing dissections via angiography inherently carries subjectivity, as numerous subtle dissections may escape detection in angiographic images. Therefore, future research should employ more detailed and standardized evaluations using intravascular imaging techniques such as IVUS and OCT. Secondly, the limited sample size, single-center design, and brief follow-up period in our study highlight the need for more extensive, multicenter, and prolonged studies to validate the effects of oral rivaroxaban on dissections post-DCB intervention, including its long-term safety. Moreover, both mechanistic studies and clinical trials are warranted to elucidate the underlying mechanisms and clinical potential of rivaroxaban in treating coronary heart disease.

## CONCLUSIONS

This study demonstrates that the combination of rivaroxaban with dual antiplatelet therapy shows promise in reducing stenosis and enhancing late lumen expansion in lesions with dissections post-DCB intervention. However, additional studies are essential to corroborate these results and assess the long-term outcomes and safety of this combined therapeutic approach.

## Data Availability

The data are not publicly available due to their containing information that could compromise the privacy of research participants.

## Abbreviations

DCB: drug-coated balloon angioplasty
ACS: acute coronary syndrome
BA: balloon angioplasty
LLL: late lumen loss

## Notes

**Funding**: This work was supported by National Key Research and Development Program of China (2022YFC3602400,2022YFC3602404), Henan Medical Science and Technology Public Relations Joint Construction Project (LHGJ20220108), National Natural Science Foundation of China (82270474). The funding source did not influence any part of the submitted work, including the study design, collection, analysis, interpretation of data, the writing of the article, or decision to submit for publication.

### Competing Interest Statement

The authors have declared no competing interest.

### Clinical Trial

NCT05750758

### Funding Statement

This work was supported by National Key Research and Development Program of China (2022YFC3602400,2022YFC3602404), Henan Medical Science and Technology Public Relations Joint Construction Project (LHGJ20220108), National Natural Science Foundation of China (82270474). The funding source did not influence any part of the submitted work, including the study design, collection, analysis, interpretation of data, the writing of the article, or decision to submit for publication.

### Author Declarations

The trial followed a prospective, randomized design. It complied with the Declaration of Helsinki and was approved by the New Business and Technology Ethics Committee of Fuwai Central China Cardiovascular Hospital, Zhengzhou University (Zhengzhou, China). All participants signed an informed consent form. This study is registered with ClinicalTrials.gov, number NCT05750758.

### Summary of Updates

Figure 1 revised; Table 1 revised;

